# Associations of Objective and Self-Reported Physical Activity and Sedentary Behavior with Skeletal Muscle Energetics: The Study of Muscle, Mobility and Aging (SOMMA)

**DOI:** 10.1101/2023.11.05.23298134

**Authors:** Yujia (Susanna) Qiao, Terri L. Blackwell, Peggy M. Cawthon, Paul M. Coen, Steven R. Cummings, Giovanna Distefano, Samaneh Farsijani, Daniel E. Forman, Bret H. Goodpaster, Stephen B. Kritchevsky, Theresa Mau, Frederico G.S. Toledo, Anne B. Newman, Nancy W. Glynn

## Abstract

**Background:** Skeletal muscle energetics decline with age, and physical activity (PA) has been shown to counteract these declines in older adults. Yet, many studies were based on self-reported PA or structured exercise interventions. We examined the associations of objective daily PA and sedentary behavior (SB) with skeletal muscle energetics and also compared with self-reported PA and SB. We also explored the extent to which PA would attenuate the associations of age with muscle energetics.

**Methods:** Among the Study of Muscle, Mobility and Aging (SOMMA) enrolled older adults, 810 (mean age=76±5, 58% women) had maximal muscle oxidative capacity measured *ex vivo* via high-resolution respirometry of permeabilized myofibers (maxOXPHOS) and *in vivo* by ^31^Phosphorus magnetic resonance spectroscopy (ATP_max_). Objective PA was measured using the wrist-worn ActiGraph GT9X over 7-days to capture sedentary behavior (SB), light, and moderate-to-vigorous PA (MVPA). Self-reported SB, MVPA, and all exercise-related PA were assessed with The Community Healthy Activities Model Program for Seniors questionnaire. Linear regression models with progressive covariate adjustments evaluated the associations between SB, PA and muscle energetics, and the attenuation of the age / muscle energetic association by PA.

**Results:** Every 30 minutes more objective MVPA was associated with 0.65 pmol/s*mg higher maxOXPHOS and 0.012 mM/sec higher ATP_max_, after adjustment for age, site/technician and sex. More time spent in objective light+MVPA was significantly associated with higher ATP_max_, but not maxOXPHOS. In contrast, every 30 minutes spent in objective SB was associated with 0.43 pmol/s*mg lower maxOXPHOS and 0.004 mM/sec lower ATP_max_. Only associations with ATP_max_ held after further adjusting for socioeconomic status, body mass index, lifestyle factors and multimorbidities. Self-reported MVPA and all exercise-related activities, but not SB, yielded similar associations with maxOXPHOS and ATP_max_. Lastly, age was only significantly associated with muscle energetics in men. Adjusting for objective time spent in MVPA attenuated the age association with ATP_max_ by nearly 60% in men.

**Conclusion:** More time spent in daily PA, especially MVPA, were associated with higher muscle energetics. Interventions that increase higher intensity activity might offer potential therapeutic interventions to slow the age-related decline in muscle energetics. Our work also emphasizes the importance of taking PA into consideration when evaluating associations related to skeletal muscle energetics.

## 1. Introduction

Skeletal muscle energetics decline with age, especially for oxidative phosphorylation (OXPHOS) and adenosine triphosphate (ATP) generation in mitochondria.^1,2^ As the key regulator of energy, mitochondria determine daily total energy production and impact skeletal muscle metabolism and quality,^3,4^ as well as metabolic, cardiovascular, renal, and neurodegenerative diseases,^5,6^ and immune responses.^7^ Importantly, lower skeletal muscle energetics have been associated with worse mobility and poor cardiorespiratory fitness in older adults, ^8–12^ which may further limit their physical activity levels. Some exercise intervention studies in older adults have shown that structured and/or supervised exercise improves skeletal muscle energetics.^13,14^ Meanwhile, daily physical activity has also been suggested as beneficial to attenuate age-related decline in mitochondrial function, yet most observational studies relied on self-reported physical activity which has been shown prone to recall bias. ^13,15–17^

Recently, a study from the Baltimore Longitudinal of Aging (BLSA) found that objectively measured moderate-to-vigorous physical activity (MVPA) was strongly associated with muscle oxidative capacity measured *in vivo* among adults aged 22 to 92 years old.^18^ Another small study also showed that maximal OXPHOS measured *ex vivo* was correlated with objectively measured daily step count using activPAL3 monitor.^19^ However, more comprehensive characterization of daily physical activity intensity levels and sedentary behavior using objective measures and examining their associations with skeletal muscle energetics are warranted to better inform optimal modality, intensity and duration of activity aiming to ameliorate age-related skeletal muscle energetics decline. In addition, comparing objective and self-reported physical activity and sedentary behavior in relation to skeletal muscle energetics in a large population of older adults will advance our knowledge base and aid interpretation of future research findings.

Furthermore, it is not clear whether age-related decline in skeletal muscle energetics is attributable to aging per se or whether it could be largely explained by concurrent decline in physical activity with aging.^20–22^. Thus, using data from older adults aged ≥70 years enrolled in the Study of Muscle, Mobility and Aging (SOMMA), we examined the associations of objectively measured physical activity intensity levels and sedentary behavior with two measures of skeletal muscle energetics (i.e., maximal OXPHOS and maximal ATP production). We further compared the aforementioned associations with a self-reported physical activity measure. Lastly, we explored to what extent physical activity could attenuate the associations of age with skeletal muscle energetics.

## 2. Material and methods

### 2.1 Study Sample

SOMMA (http://sommaonline.ucsf.edu) is a prospective longitudinal on-going cohort study to examine muscle properties that most strongly predict mobility disability and decline in fitness as people age. From April 2019 through December 2021, 879 community-dwelling older adults (≥70 years) with a 4-meter gait speed of ≥0.6 m/s and willing/eligible to have magnetic resonance (MR) spectroscopy and a skeletal muscle biopsy were enrolled in SOMMA at two academic clinical centers (University of Pittsburgh and Wake Forest University School of Medicine). Participants were excluded if they reported an inability to walk one-quarter of a mile or climb a flight of stairs; had body mass index (BMI) ≥40 kg/m^2^; had an active malignancy or dementia; or any medical contraindication to biopsy or MR. At baseline, participants completed three clinic visits within six weeks of each other. Details of the study design and timing of clinical measurements can be found elsewhere.^23^ All participants provided written informed consent, and SOMMA was approved by the WIRB-Copernicus Group Institutional Review Board (WCG IRB) as the single IRB for all participating sites.

### 2.2 Objectively Measured Physical Activity

The ActiGraph GT9X Link (ActiGraph, LLC, Pensacola, FL, USA) is a validated 3-axial accelerometer providing objective measurements of movement.^24^ At the first of three baseline clinic exams, participants were instructed to wear the device on their non-dominant wrist for at least 7 consecutive full days at all times except under water for more than 30 minutes.

Data were collected at 80 hertz. Non-wear time was determined using Troiano 2007 non-wear algorithm.^25^ Trained staff further examined the data marked as non-wear and verified non-wear time after reviewing results from the off-wrist sensor in the device.^26^ A valid day was defined as ≥17 hours wear time during a 24-hour period (00:00 to 23:59 for the same date). The first day of wear was excluded from these analyses, as the participants were required to do a number of physical performance tests during their clinic visit and the activity level may not be representative of their usual activity patterns. Activity level was captured in one-minute epochs with vector magnitude counts (VMC), a measure that combines data from all 3 axes. For non-wear time, VMC was imputed by replacing the missing value with the average activity for that minute across all days of wear.^27^ For example, if data was missing on one day for 10:39AM, then the average of the VMC at 10:39AM on all other days was used. Of all 24-hr intervals across all participants, 97% had no missing data. To better represent older adults’ physical activity intensities measured with a wrist-worn device, we used cutpoints from Montoye et al.^28^ based on VMC to classify sum of total time spent (min/day) in sedentary behavior (<2860 VMC/min), light+MPVA (≥2860 VMC/min) and moderate-to-vigorous physical activity (MVPA, ≥3941 VMC/min). For sedentary time, we excluded the time the participant was in bed. Daily activity was also assessed as total activity count per 24-hour day to avoid potential error and bias that may be introduced by converting the data to energy expenditure using population level equations.^27^ Activity parameters were averaged over valid days to obtain a more representative characterization of usual activity patterns.^29^

In addition, participants also wore the activPAL4™ (PAL Technologies Ltd, Glasgow, Scotland) on their right thigh concurrent with wearing the ActiGraph. Data were collected at 20 hertz and proprietary algorithms use the accelerometer measurements to determine posture (i.e. sitting, lying, standing) and calculated daily step count. The sample size for participants with step count data was smaller (n=713) due to technical problems, thus we examined the associations between daily step count and muscle energetics as a sensitivity analysis.

### 2.3 Self-reported Physical Activity

The Community Healthy Activities Model Program for Seniors (CHAMPS) is a commonly-used and validated 40-item questionnaire to assess physical activity levels in older adults.^30^ The CHAMPS questionnaire queries activities performed retrospectively from sedentary behavior (e.g., reading, attending a concert, attending church, attending group meetings, using a computer, playing cards or board games), light (e.g., leisurely walking, light work around the house, going to the senior center) to more moderate-to-vigorous intensities (e.g., jogging or running, brisk walking, basketball, moderate to heavy strength training) in the past 4 weeks. Each activity was classified into one of three physical activity intensity levels using metabolic equivalents (METs): sedentary = 1 METs, light = METs >1 and <3, MVPA = METs ≥3.^31^ To better compare with SOMMA accelerometry data, we used the following derived summary metrics from the CHAMPS: sum of total time spent (min/day) in sedentary behavior (METs ≤1), moderate-to-vigorous physical activity (MVPA, METs ≥3), and all exercise-related physical activities (METs ≥2).

### 2.4 Skeletal Muscle Energetics

A skeletal muscle biopsy was obtained from the vastus lateralis after 12-hour fast and limited strenuous exercise for 48 hours before the procedure, collected within six weeks of the first SOMMA baseline visit. Participants also rested in a supine position for at least 15 minutes prior to the biopsy. The biopsy specimen was collected ∼15cm above the patella using a 5 or 6mm Bergstrom-style core biopsy needle with suction under local anesthesia (1% or 2% lidocaine). Details on muscle tissue and fiber bundle preparation have been published.^12^

Complex I- and II-supported maximal OXPHOS (maxOXPHOS), also known as state 3 respiration, was assessed *ex vivo* with high-resolution respirometry. Permeabilized fiber bundles were placed into the respirometer chambers of an Oxygraph-2K (O2K, Oroboros Instruments, Austria). MaxOXPHOS was measured in the presence of saturated concentrations of pyruvate, malate, ADP, glutamate and succinate. Measurements were performed in duplicate at 37°C. Integrity of the outer mitochondrial membrane was tested with Cytochrome *c*, and any sample with a response greater than 15% was omitted from analysis. Steady-state oxygen flux was normalized to wet weight of the fiber bundle using Datlab 7.4 software. In SOMMA, the mean coefficient of variation for duplicates of maxOXPHOS measurement was 11.5% across both clinical sites.^12^

Maximal ATP production (ATP_max_) was quantified by ^31^P MRS in a 3 Tesla MR scanner (Siemen’s Medical System—Prisma at Pittsburgh site or Skyra at Wake Forest site). This is a functional measure of *in-vivo* ATP production, using the rate of regeneration of phosphocreatine (PCr) after a short bout of exercise.^8^ Participants were instructed to lie in a supine position with their lower leg strapped and right knee joint in 20°-30° of flexion. A 12’’ dual-tuned, surface RF coil (PulseTeq, Limited) was placed over the right distal vastus lateralis. Participants did two bouts of isometric knee extension against the resistance of an ankle strap.^32^ PCr recovery rate after exercise until PCr returned to baseline levels was fit and the time-constant of the mono-exponential fit (tau) was used to calculate ATP_max_^.33,34^ In SOMMA, the mean coefficient of variation for duplicates of ATP_max_ measurement was 9.9% across both clinic sites.^12^

### 2.5 Covariates

Age, sex, race (White and non-white [including Black, Asian, Native American/Alaskan Native, Native Hawaiian/Pacific Islander, Multi-Racial, Unknown]), and education (high school or less, some college, college graduate, post college work) were self-reported. Smoking status (current/former/never), and alcohol intake (drinks/week) were also recorded via questionnaire. Height without shoes and weight with light clothing were measured, and further used to calculate body mass index (BMI, kg/m^2^). We queried self-report of a physician diagnosis history (yes/no) of several health conditions. A composite multimorbidity index score was calculated using a modified list of chronic conditions from the Rochester Epidemiology Project.^35^ The score included 11 age-related conditions: cancer, chronic kidney disease or renal failure, atrial fibrillation, lung disease (i.e., chronic obstructive pulmonary disease, bronchitis, asthma, or emphysema), coronary heart disease (i.e., blocked artery or myocardial infarction), depression (defined as ≥10 score on the 10-item Center for Epidemiologic Studies Depression Scale^39^), heart failure, dementia, diabetes, stroke, and aortic stenosis.^36^ Number of prescription medications (Rx) was confirmed by participants bringing all medication containers they had taken in the 30 days prior to any of the 3 visits; alternatively, clinic staff obtained this information over the telephone or at a return visit. Values for number Rx medications were truncated to 20 (n=2 with >20 Rx medications).

### 2.6 Statistical Analyses

Descriptive characteristics were reported as mean ± standard deviation (SD) or frequencies (percentages) overall and by sex for the final analytical sample with physical activity measures and ≥1 muscle energetics (N=810); sample size varied based on missingness for skeletal muscle energetics (n=698 for maxOXPHOS, n=755 for ATP_max_, Supplemental Figure 1). Comparisons by sex were evaluated using two-sample t tests for normally distributed continuous variables, Wilcoxon rank-sum test for skewed continuous variables, and a chi-square test for categorical variables or a Fisher’s exact test for those with low expected cell counts.

To evaluate the associations between physical activity and skeletal muscle energetics, we generated linear regression models with progressive covariate adjustments to account for established and potential confounders. All physical activity metrics were scaled to every 30 min/day, except for the accelerometer-measured total activity count in 24 hours. Model 1 adjusted for technician/site, age, and sex; Model 2 further adjusted for race, education, BMI, smoking status, alcohol intake, multimorbidity index, and number of Rx. Each physical activity metric was modeled separately in relation to maxOXPHOS and ATP_max_. Furthermore, to assess the effect of physical activity on the associations of age with muscle energetics, we also generated crude models with only technician/site, sex, and age, with muscle energetics as dependent variables; and compared them to adjusted model 1 to obtain the percentage of attenuation in coefficients of age after adjusting for physical activity.

All analyses were conducted in the overall sample and stratified by sex. We tested for sex interactions with all physical activity variables by modeling the main effects (e.g., sex and time spent in MVPA) and their product (e.g., sex*time spent in MVPA) together adjusting for technician/site and age. We also conducted a sensitivity analysis using the objective physical activity metric of daily step count and examined its association with skeletal muscle energetics.

Alpha was set to 0.05 and two-sided P values smaller than 0.05 were considered significant. All analyses were performed using SAS software, version 9.4 (SAS Institute, Inc, Cary, NC).

## 3. RESULTS

### 3.1 Participant Characteristics

Participants (N=810) were 76.4 ± 5.0 years old (range: 70-94), with 58% women, 85% white and 86.2% having some college or higher education (Table 1). Their average BMI was 27.6 ± 4.6 kg/m^2^ with no sex difference (Table 1). On average, men reported fewer miltimorbidities than women, with had higher prevalence of heart disease and diabetes mellitus (Table 1). Men had higher values for maxOXPHOS and ATP_max_ compared to women (both P <0.001) (Table 1).

**Table 1.** Baseline characteristics of the full sample and stratified by sex in the Study of Muscle, Mobility and Aging (SOMMA)

| Characteristic | All<br>(N= 810) | Men<br>(n= 337) | Women<br>(n= 473) | P-value |
| --- | --- | --- | --- | --- |
| Age, years | 76.4 ± 5.0 | 76.2 ± 5.1 | 76.5 ± 5.0 | 0.514 |
| Race |  |  |  | 0.111 |
| White | 690 (85.2) | 295 (87.5) | 395 (83.5) |  |
| Nonwhite | 120 (14.8) | 42 (12.5) | 78 (16.5) |  |
| College education |  |  |  | <.001 |
| High school or less or other | 111 (13.8) | 39 (11.7) | 72 (15.4) |  |
| Some college | 190 (23.7) | 49 (14.7) | 141 (30.1) |  |
| College graduate | 208 (25.9) | 111 (33.2) | 97 (20.7) |  |
| Post college work | 293 (36.5) | 135 (40.4) | 158 (33.8) |  |
| Body mass index, kg/m<sup>2</sup> | 27.6 ± 4.6 | 27.9 ± 4.2 | 27.4 ± 4.9 | 0.153 |
| Smoking status |  |  |  | 0.614 |
| Never | 450 (55.9) | 182 (54.5) | 268 (56.9) |  |
| Past | 330 (41.0) | 143 (42.8) | 187 (39.7) |  |
| Current | 25 (3.1) | 9 (2.7) | 16 (3.4) |  |
| Alcohol intake, drinks/week | 2.8 ± 4.6 | 3.5 ± 5.1 | 2.3 ± 4.1 | <.001 |
| SOMMA Multimorbidity Index (0-11) | 0.8 ± 0.9 | 0.9 ± 0.9 | 0.7 ± 0.8 | 0.006 |
| History of cancer<sup>a</sup> | 204 (25.3) | 95 (28.4) | 109 (23.1) | 0.090 |
| History of cardiac arrhythmia | 32 (4.0) | 13 (3.9) | 19 (4.0) | 0.917 |
| History of kidney disease or renal failure | 30 (3.7) | 12 (3.6) | 18 (3.8) | 0.871 |
| History of COPD | 107 (13.3) | 37 (11.0) | 70 (14.8) | 0.118 |
| History of coronary heart disease<sup>b</sup> | 59 (7.3) | 47 (14.0) | 12 (2.5) | <.001 |
| History of congestive heart failure | 6 (0.7) | 5 (1.5) | 1 (0.2) | 0.087 |
| Dementia | 0 | 0 | 0 | N/A |
| Depression<sup>c</sup> | 63 (7.9) | 22 (6.7) | 41 (8.8) | 0.280 |
| History of diabetes mellitus | 126 (15.6) | 63 (18.8) | 63 (13.3) | 0.035 |
| History of stroke | 21 (2.6) | 11 (3.3) | 10 (2.1) | 0.306 |
| History of aortic stenosis | 7 (0.9) | 2 (0.6) | 5 (1.1) | 0.706 |
| Number of Rx medications taken in the last 30 days | 4.4 ± 3.5 | 4.2 ± 3.3 | 4.6 ± 3.6 | 0.210 |
| Skeletal muscle energetic measures: |  |  |  |  |
| maxOXPHOS, pmol/(s*mg) | 59.5 ± 18.4 | 65.4 ± 19.7 | 54.8 ± 15.9 | <.001 |
| ATP<sub>max</sub>, mM/sec | 0.54 ± 0.15 | 0.57 ± 0.16 | 0.52 ± 0.13 | <.001 |
| Objective activity metrics: |  |  |  |  |
| Sedentary behavior<sup>d</sup>, min/day | 676.8 ± 107.6 | 710.3 ± 97.4 | 653.0 ± 108.2 | <.001 |
| MVPA<sup>d</sup>, min/day | 186.6 ± 86.0 | 161.3 ± 74.1 | 204.6 ± 89.4 | <.001 |
| Light+MVPA<sup>d</sup>, min/day | 284.3 ± 100.7 | 253.3 ± 88.1 | 306.4 ± 103.4 | <.001 |
| Total activity counts<sup>d</sup>, per 10000 counts/day | 198 ± 59 | 182 ± 52 | 210 ± 60 | <.001 |
| Daily step count<sup>e</sup>, steps/day | 6860.5 ± 3204.0 | 7214.9 ± 3499.5 | 6608.3 ± 2954.4 | 0.016 |
| Self-reported activity metrics: |  |  |  |  |
| Sedentary behavior, min/day | 154.7 ± 60.7 | 147.0 ± 62.8 | 160.1 ± 58.7 | 0.003 |
| MVPA (METs ≥3), min/day | 59.5 ± 58.2 | 71.9 ± 64.4 | 50.7 ± 51.7 | <.001 |
| All exercise-related activities (METs ≥2), min/day | 129.0 ± 88.7 | 135.9 ± 95.1 | 124.1 ± 83.6 | 0.190 |

Note: All reported in mean ± SD or n (%).
<sup>a</sup> History of cancer excluded nonmelanoma skin cancer.
<sup>b</sup> History of heart disease included blocked artery or myocardial infraction.
<sup>c</sup> Depression was defined as Center for Epidemiologic Studies Depression Scale ≥10.
<sup>d</sup> ActiGraph GT9X data were averaged over all 24 hr periods of wear (00:00 to 23:59). Sedentary behavior excluded time in-bed.
Physical activity intensities were defined using Montoye et al. 2020 based on vector magnitude counts (VMC) to classify sum of total time spent (min/day) in sedentary behavior (<2860 VMC/min), light activity (2860-3940 VMC/min) and MVPA (≥3941 VMC/min).
<sup>e</sup> activePAL data were averaged over all 24 hr periods of wear (00:00 to 23:59).
Abbreviations: COPD = chronic obstructive lung disease; OXPHOS = oxidative phosphorylation; ATP = adenosine triphosphate; CHAMPS = Community Healthy Activities Model Program for Seniors; MVPA = moderate-to-vigorous physical activity; METs = metabolic equivalents.

Based on objective PA data, participants spent almost 50% of their time being sedentary (excluding sleep), and nearly 13% (3.1 ± 1.4 hour) spent in MVPA (Table 1). Interestingly, when compared to women, men had ∼9% more minutes spent in objective sedentary behavior and ∼27% less in objective MVPA (Table 1). The total activity counts/day also confirmed that women were more active overall throughout the day than men (P <0.001, Table 1). However, women had a lower daily step count than men (P=0.016, Table 1). When evaluating activity levels using self-reported compared to objective data, participants self-reported four times fewer minutes spent in sedentary behavior and three times fewer minutes of MVPA (Table 1). Counter to the data for objectively measured PA, men self-reported ∼8% less minutes spent in sedentary behavior and ∼41% more time spent in MVPA as compared to women. Overall, the correlations between objective and self-reported measures were low (Sedentary time r=0.03, MVPA r=0.25).

### 3.2 Associations between Objectively Measured Physical Activity, Sedentary Behavior and Skeletal Muscle Energetics

Overall, higher MVPA was more strongly associated with both maxOXPHOS and ATP_max_ than sedentary behavior. Specifically, every 30 minutes more in MVPA was associated with 0.65 (95% CI: 0.17, 1.13) higher pmol/(s*mg) maxOXPHOS and 0.012 (95% CI: 0.009, 0.016) higher mM/sec ATP_max_, after minimal adjustment (Table 2, Model 1). Time spent in light+MVPA was only significantly associated with ATP_max_. Furthermore, every 30 minutes more in sedentary behavior was associated with 0.39 (95% CI: −0.77, −0.02) lower pmol/(s*mg) maxOXPHOS and 0.006 (95% CI: −0.009, −0.003) lower mM/sec ATP_max_ (Table 2, Model 1). However, the associations with maxOXPHOS, but not with ATP_max_, were fully attenuated after further adjustment for socioeconomic status, BMI, lifestyle factors and multimorbidity index (Table 2, Model 2).

**Table 2.** Associations of physical activity and skeletal muscle energetics in the Study of Muscle, Mobility and Aging (SOMMA)^a^.

| Physical activity measures | maxOXPHOS (pmol/(s*mg)) |  | ATP<sub>max</sub> (mM/sec) |  |
| --- | --- | --- | --- | --- |
|  | Model 1 | Model 2 | Model 1 | Model 2 |
|  | β (95%CI) | β (95%CI) | B (95%CI) | β (95%CI) |
| Objective metrics<sup>b</sup>: |  |  |  |  |
| Sedentary behavior<sup>c</sup> | **-0.39**<br>(-0.77, -0.02) | -0.05<br>(-0.44, 0.35) | **-0.006**<br>(-0.009, -0.003) | **-0.005**<br>(-0.008, -0.002) |
| Light+MVPA<sup>c</sup> | 0.38<br>(-0.02, 0.79) | 0.003<br>(-0.42, 0.43) | **0.010**<br>(0.006, 0.013) | **0.008**<br>(0.005, 0.012) |
| MVPA<sup>c</sup> | **0.65**<br>(0.17, 1.13) | 0.15<br>(-0.35, 0.66) | **0.012**<br>(0.009, 0.016) | **0.011**<br>(0.007, 0.015) |
| Total activity count<sup>d</sup> | **1.90**<br>(0.54, 3.26) | 0.65<br>(-0.77, 2.07) | **0.037**<br>(0.026, 0.047) | **0.033**<br>(0.022, 0.044) |
| Self-reported metrics: |  |  |  |  |
| Sedentary behavior<sup>c</sup> | 0.43<br>(-0.22, 1.07) | -0.08<br>(-0.74, 0.58) | 0.004<br>(-0.002, 0.009) | 0.001<br>(-0.004, 0.006) |
| All exercise-related (METs ≥2)<sup>c</sup> | **0.92**<br>(0.49, 1.35) | **0.53**<br>(0.09, 0.97) | **0.011**<br>(0.008, 0.015) | **0.009**<br>(0.005, 0.013) |
| MVPA (METs ≥3)<sup>c</sup> | **1.88**<br>(1.23, 2.53) | **1.24**<br>(0.57, 1.92) | **0.022**<br>(0.016, 0.027) | **0.019**<br>(0.013, 0.024) |

Note: Bold β were significant as P<0.05.
<sup>a</sup> The analytical samples were n=697 for maxOXPHOS, n=754 for ATP<sub>max</sub>. Model 1 adjusted for age, sex, technician/site. Model 2 further adjusted for race, education, body mass index, smoking, alcohol, number of comorbidities, number of Rx medications.
<sup>b</sup> ActiGraph GT9X data were averaged over all 24 hr periods of wear (00:00 to 23:59). Sedentary behavior excluded time in-bed. Physical activity intensities were defined using Montoye et al. 2020 based on vector magnitude counts (VMC) to classify sum of total time spent (min/day) in sedentary behavior (<2860 VMC/min), light activity (2860-3940 VMC/min) and MVPA (≥3941 VMC/min).
<sup>c</sup> Re-scaled to every 30 min/day more.
<sup>d</sup> Re-scaled to 1 standard deviation = 585821 count/day more.
Abbreviations: OXPHOS = oxidative phosphorylation; ATP = adenosine triphosphate; CHAMPS = Community Healthy Activities Model Program for Seniors; MVPA = moderate-to-vigorous physical activity; METs = metabolic equivalents.

Differential associations were also observed after sex stratification. Notably, there were no significant interactions with maxOXPHOS (Supplemental Table 1); whereas the associations with ATP_max_ were stronger in men than women across all physical activity intensities, except for sedentary behavior (all P_sex-interaction_ <0.01) (Supplemental Table 2).

In the sensitivity analysis using the objective measure of daily step count, every 1000 steps/day more was associated with 0.97 (95% CI: 0.54, 1.41) higher pmol/(s*mg) maxOXPHOS and 0.016 (95% CI: 0.013, 0.020) higher mM/sec ATP_max_, after fully adjustment (Supplemental Table 3). No sex difference was found (all P_sex-interaction_ >0.3) (Supplemental Table 3).

### 3.3 Associations between Self-reported Physical Activity, Sedentary Behavior and Skeletal Muscle Energetics

Overall, time spent in MVPA, and all exercise-related activity had almost 2 times greater magnitude of the associations with skeletal muscle energetics than objective PA, while self-reported sedentary behavior was not associated with any skeletal muscle energetics (Table 2). After full adjustment, every 30 minutes more in MVPA and all exercise-related activities were associated with 1.24 (95% CI: 0.57, 1.92) and 0.53 (95%CI: 0.09, 0.97) higher pmol/(s*mg) maxOXPHOS, respectively; and 0.019 (95% CI: 0.013, 0.024) and 0.009 (95% CI: 0.005, 0.013) higher mM/sec ATP_max_, respectively (Table 2, Model 2). Furthermore, these associations for ATP_max_ were stronger in women than men when stratified by sex (all P_sex-interaction_ ≤0.08) (Supplemental Table 1).

### 3.4 Associations of Age with Muscle Energetics

In the overall sample, every 5-years older in age was associated with 1.72 (95% CI: - 2.99, −0.46) pmol/(s*mg) and 0.015 (95% CI: −0.025, −0.005) mM/sec lower maxOXPHOS and ATP_max_, respectively (Figure 1). The associations of age with skeletal muscle energetics were significant in men but not women. Notably, among men, adjusting for objective MVPA or sedentary behavior had little impact the associations of age with maxOXPHOS (Figure 1 (A)). Yet, time spent in objective MVPA attenuated the age association with ATP_max_ among men by 58% (Figure 1 (B)).

**Figure 1.**
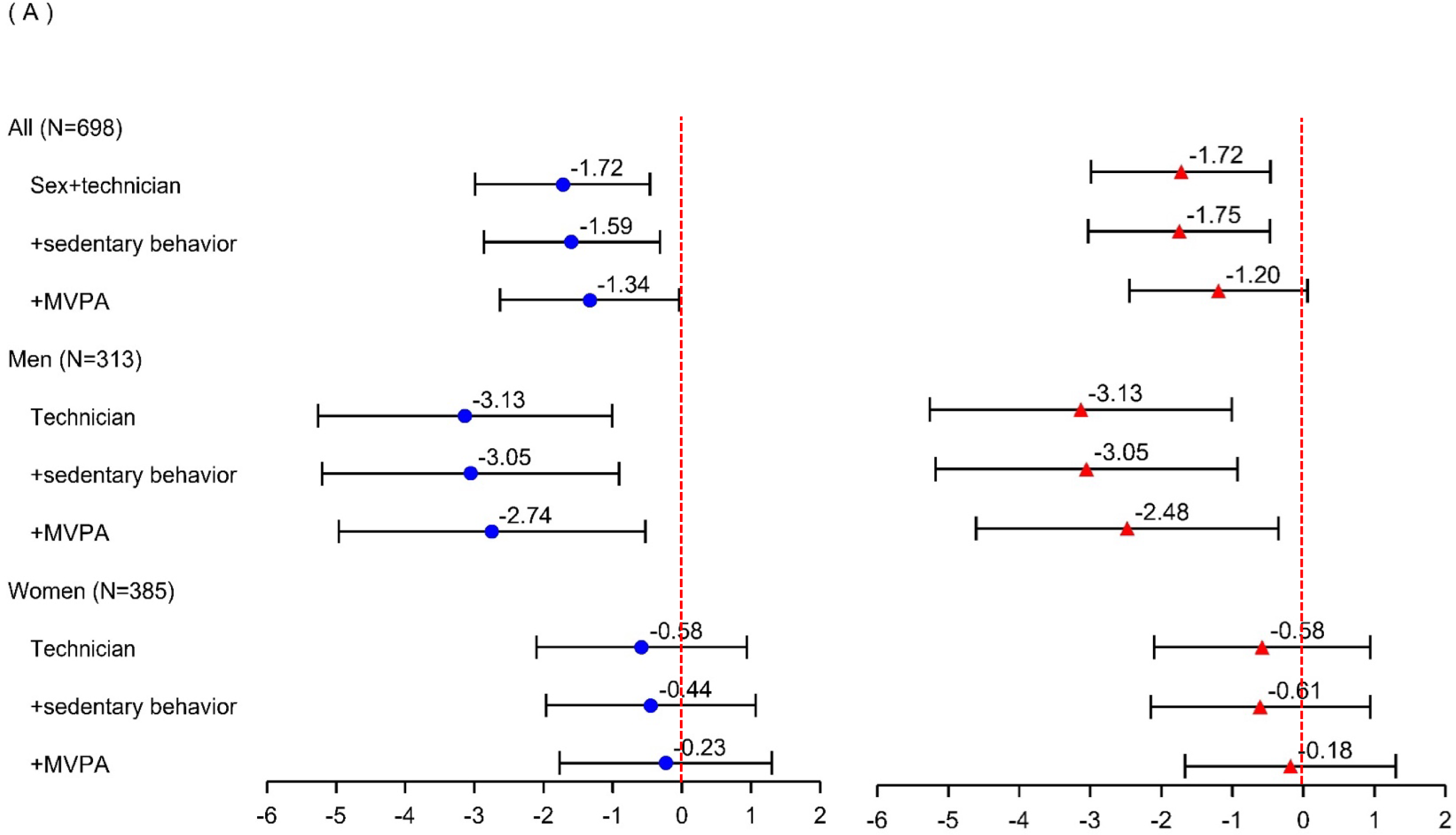
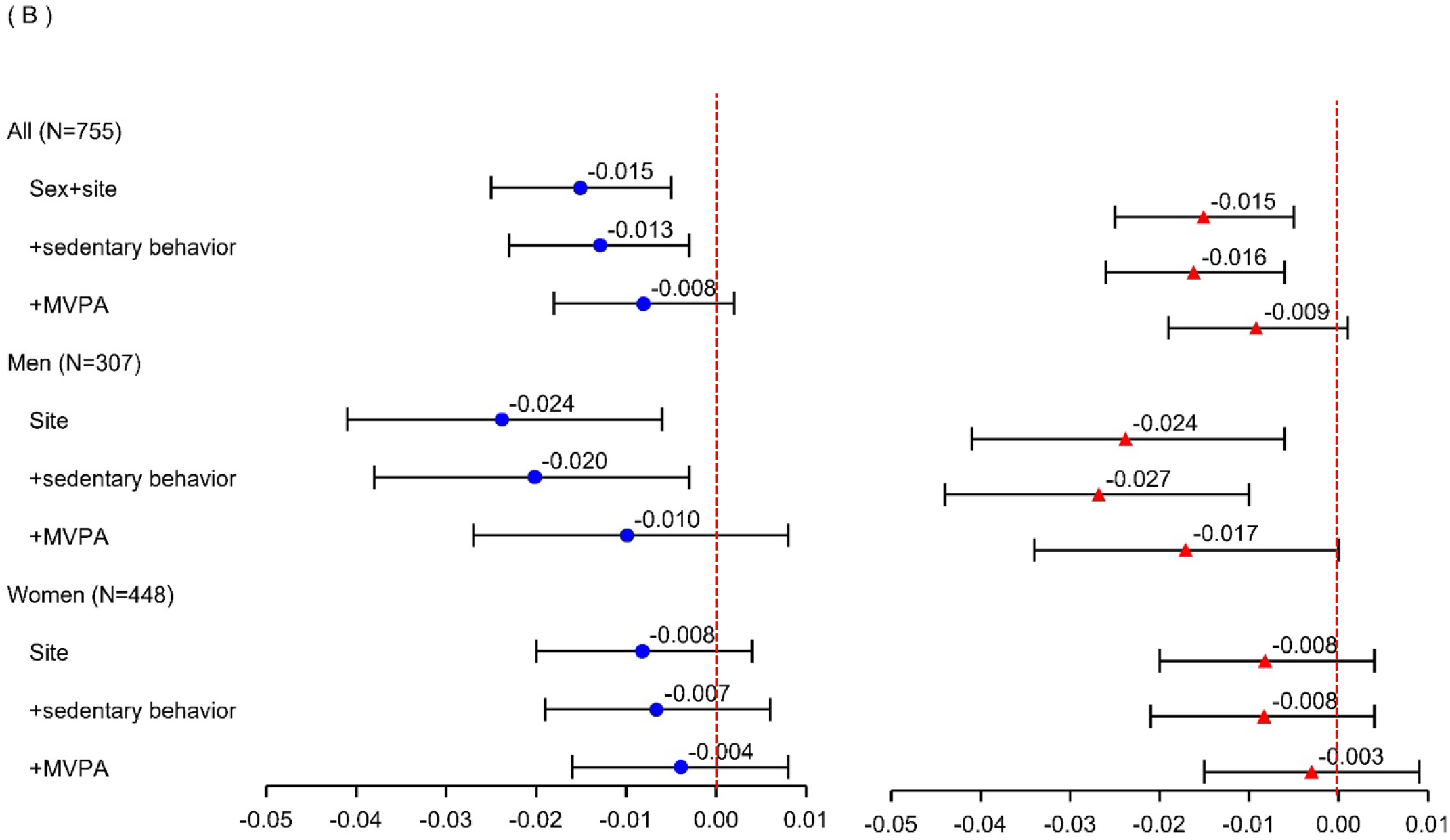
Associations of age per 5-year older and skeletal muscle energetics with physical activity in the Study of Muscle, Mobility and Aging (SOMMA). Panel (A) is for maxOXPHOS, panel (B) is for ATP_max_. Ojective data are shown in blue circles, self-report as red triangles. All reported in β (95%CI). ActiGraph GT9X data were averaged over all 24 hr periods of wear (00:00 to 23:59). Physical activity intensities were defined using Montoye et al. 2020 based on vector magnitude counts (VMC) to classify sum of total time spent (min/day) in sedentary behavior (<2860 VMC/min), light activity (2860-3940 VMC/min) and MVPA (≥3941 VMC/min). Abbreviations: OXPHOS = oxidative phosphorylation; ATP = adenosine triphosphate; CHAMPS = The Community Healthy Activities Model Program for Seniors; MVPA = moderate-to-vigorous physical activity; METs = metabolic equivalents.

## 4. DISCUSSION

Those with greater time spend in spent daily physical activity, particularly higher intensity (i.e., MVPA), had significantly greater skeletal muscle energetics, while more time spent in daily sedentary behavior was associated with lower skeletal muscle energetics. In addition, we found sex differences in associations between physical activity and ATP_max_. Age was associated with skeletal muscle energetics in men but not women. Adjusting for time spent in either objective or self-reported MVPA attenuated coefficient of age by ∼60%, fully explaining the association between age and ATP_max_. Yet, the association between age and maxOXPHOS held after adjusting for either objective or self-reported MVPA. Thus, our findings might indicate that higher intensity physical activity could offset the age-related decline in ATP_max_ or that lower ATP_max_ impedes physical activity at older ages.

Both objective and self-reported physical activity revealed that those who spent more time in physical activity, especially MVPA, had higher skeletal muscle energetics, concurring with previous studies.^4,15,18,22,37^ A plausible biological mechanism behind the health benefits of physical activity is neuromuscular adaptation, including muscle and myofiber hypertrophy and sizable gains in strength and power, which further induce signaling pathways to improve mitochondrial function.^14,16^ Further, some studies have suggested that only regular daily physical activity above light intensity may achieve favorable mitochondrial adaptations in muscle,^18,38^ which may explain why we observed stronger and more robust associations between time spent in MVPA and skeletal muscle energetics. Although many existing intervention studies have observed improvement in mitochondrial respiration and oxidative phosphorylation after resistance and endurance exercise training,^39–41^ further exploration of optimal frequency, intensity and duration of daily physical activity to maximize its health benefits on skeletal muscle energetics would also be informative.

Additionally, we also found that those who spent more time in objectively measured sedentary behavior had worse skeletal muscle energetics. Randomized controlled trials have found that two weeks of leg immobilization or 10 days of bed rest decreased skeletal muscle respiration in older men.^42,43^ Thus, our results align with intervention studies that show that sedentariness is associated with worse muscle energetics.

Furthermore, our findings suggested that higher intensity physical activity attenuated and fully explained the associations of age with ATP_max_. Previous studies have emphasized the importance of accounting for physical activity when examining associations related to skeletal muscle energetics. Similar to our findings, Kent-Braun and Ng^20^ showed that after adjusting for objectively measured physical activity, age was no longer associated with t_1/2_ of PCr recovery measured *in vivo* at the tibialis anterior using MRS. Whereas, Adelnia et al. found that, although accounting for objective MVPA reduced the coefficient for age by ∼40%, objective MVPA did not completely explain the age association with recovery time constant of the PCr using the same *in vivo* ^31^P MRS method,^18^ which might be due to their slightly younger age population than SOMMA. On the other hand, Larsen et al.^44^ found maximal OXPHOS measured *ex vivo* was similar in older and younger adults when matched on their physical activity and cardiorespiratory fitness level, which contradicted our observations of significant age associations with maxOXPHOS after adjusting for MVPA in SOMMA. Such differential findings might be due to different substrates and lab protocol deployed to quantify maximal OXPHOS. Furthermore, We speculate that our differential findings in terms of physical activity effect on the association of age with ATP_max_ versus maxOXPHOS center around the *in vivo* versus *ex vivo* measures. The *in vivo* ATP_max_ measure reflects more physiological factors beyond maxOXPHOS, such as cardiorespiratory fitness, oxygen delivery, muscle perfusion, and neuromuscular activation, whereas *ex vivo* maxOXPHOS may be a more direct mitochondrial respiration indicator. Differences in these protocols related to error and precision could also explain our disparate results. Prospective longitudinal data are still needed to decipher the causal relationships of physical activity on age-related decline in skeletal muscle energetics.

Notably, there were some inconsistencies in objectively measured versus self-reported versus time engaging in physical activity and sedentary behavior that may have further influenced their differential associations with skeletal muscle energetics. For MVPA, we observed a slightly higher correlation between objective and self-reported measures (r=0.25, p<0.05), yet it is still lower than previous finding showing that the CHAMPS questionnaire was moderately correlated with the objective SenseWear Armband (rho=0.42, p=0.002).^45^ In SOMMA, the objective measure classified more time spent in MVPA than by self-reported. Conversely to self-reported, the objective measure showed longer time in MVPA in women than men. Such differences could be largely driven by either the finite list of MVPA-related activities included in the CHAMPS questionnaire, or that the Montoye 2020 cut-points^28^ to classify objective MVPA in SOMMA might potentially mis-classify. We chose the Montoye cut-points to better accommodate higher energy expenditure among older adults given the same load of activity, yet those cut-points are lower than commonly used cut-points used for general adult populations, thus likely classified more time spent in MVPA. Another possible explanation is that household activities are performed across a wide spectrum of METs (e.g., vacuuming and dough kneading range from 1.5 METs to 5 METs).^47^ An objective measure is able to capture varied degree of intensity, while a self-reported questionnaire can only assign a fixed intensity level to the same activity item, for example, light work around the house equals to 2.5 METs.^30^

For sedentary behavior, we observed no significant correlation between objective and self-reported measures (r=0.03), and the objective measure classified almost 4 times more minutes in sedentary behavior than participants self-reported. In addition to potential recall bias, the CHAMPS questionnaire was not designed to capture all types of sedentary behavior or account for behavior for all 24 hours of the day; there are only 6 out of 40 items querying sedentary behavior.^30^ More descriptive data, including energy expenditure, from older adults and a better validation study for wrist-worn physical activity cut-points among older adults are needed to confirm our observation of sex difference in physical activity related to skeletal muscle energetics.

Several strengths and some limitations of our study need to be noted. First, SOMMA had extensive measures of physical activity and sedentary behaviors, using both objective and self-reported measures, providing us a unique opportunity to scrutinize the contribution of various activity measures with muscle energetics. Additionally, SOMMA had comprehensive skeletal muscle energetics measures, including both *ex vivo* maxOXPHOS and *in vivo* ATP_max_, in a large sample of older women and men ranging from 70-94 years old. Given the differences in muscle energetics across race (data not shown), our results might be limited due to the largely white sample. Yet, the values of muscle energetics measures in our study participants were comparable with the same age group demonstrated in other studies.^4,8^ Further, there might be potential residual confounding, given the complex biological pathways linking physical activity and skeletal muscle energetics. Lastly, future longitudinal work in SOMMA will be able to fully disentangle the potential bi-directionality and causality between physical activity and skeletal muscle energetics.

## 5. Conclusion

We found that higher intensity physical activity, especially MVPA, were associated with higher skeletal muscle energetics and attenuated the age effect. Future explorations of daily physical activity modalities, frequency, and intensities using objective measures could inform potential therapeutic interventions to maintain or improve skeletal muscle energetics for the advancement of longevity and healthy aging among older adults.

## Data Availability

All data produced in the present study are available upon reasonable request to the authors.

https://sommaonline.ucsf.edu

## Acknowledgements

The Study of Muscle, Mobility and Aging is supported by funding from the National Institute on Aging (AG 059416). Study infrastructure support was funded in part by NIA Claude D. Pepper Older American Independence Centers at University of Pittsburgh (P30 AG024827) and Wake Forest University (P30 AG021332) and the Clinical and Translational Science Institutes, funded by the National Center for Advancing Translational Science, at Wake Forest University (UL1 0TR001420).

## Authors’ contributions

Drs. Qiao and Glynn and Ms. Blackwell had full access to all of the data for the study and take responsibility for the integrity of the data and accuracy of the data analyses. All authors: interpretation of data, critical revision of manuscript for important intellectual content. All authors read and approved the submitted manuscript.

---

**Supplemental Table 1.**
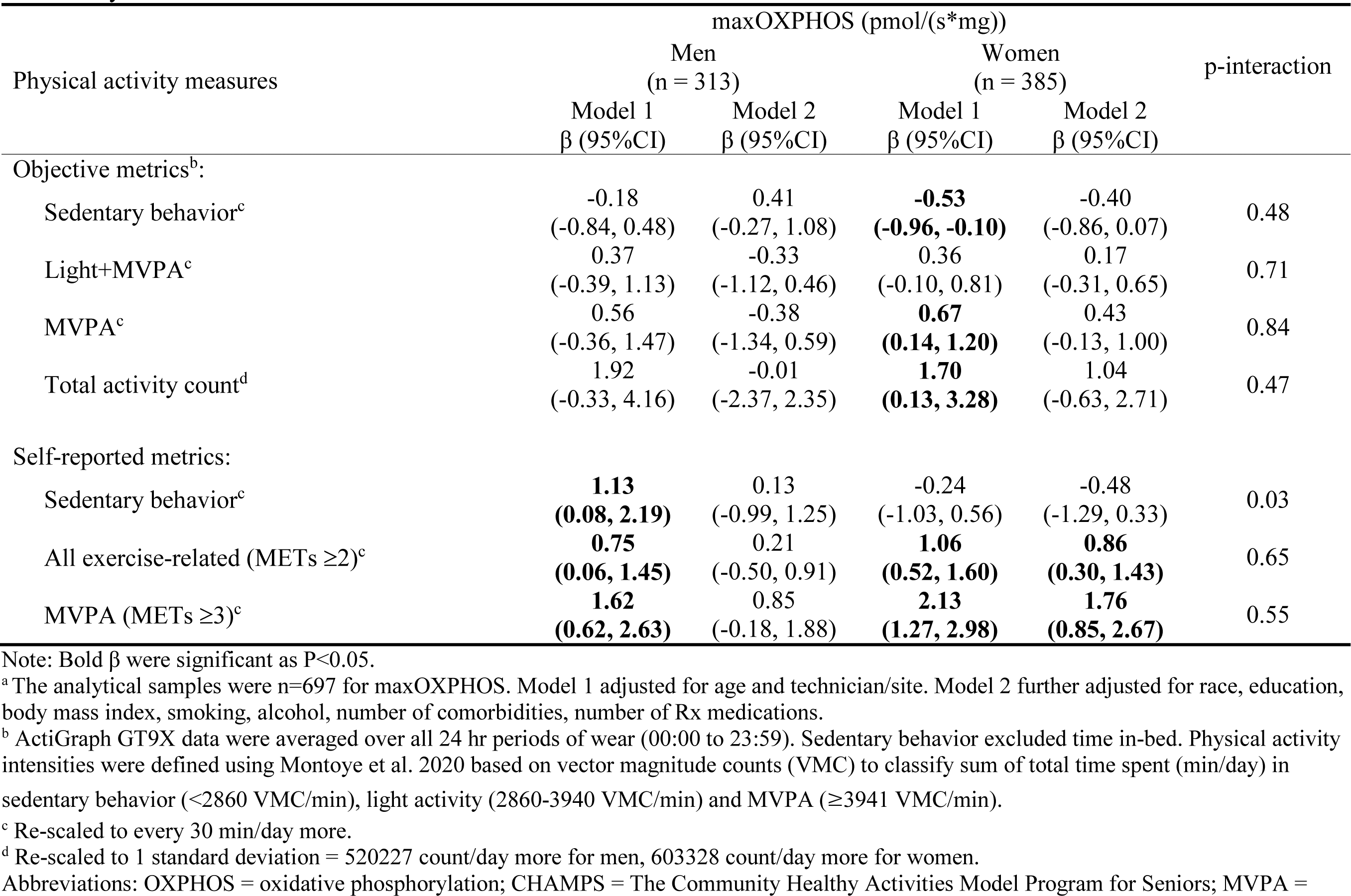
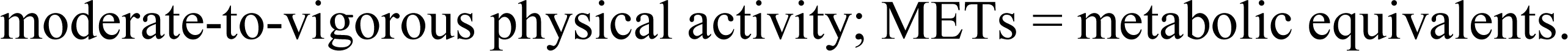
Associations of physical activity and maxOXPHOS in the Study of Muscle, Mobility and Aging (SOMMA) stratified by sex^a^.

| Physical activity measures | maxOXPHOS (pmol/(s*mg)) |  |  |  | p-interaction |
| --- | --- | --- | --- | --- | --- |
|  | Men |  | Women |  |  |
|  | (n = 313) |  | (n = 385) |  |  |
|  | Model 1 | Model 2 | Model 1 | Model 2 |  |
|  | β (95%CI) | β (95%CI) | β (95%CI) | β (95%CI) |  |
| Objective metrics<sup>b</sup>: |  |  |  |  |  |
| Sedentary behavior<sup>c</sup> | -0.18<br>(-0.84, 0.48) | 0.41<br>(-0.27, 1.08) | **-0.53**<br>**(-0.96, -0.10)** | -0.40<br>(-0.86, 0.07) | 0.48 |
| Light+MVPA<sup>c</sup> | 0.37<br>(-0.39, 1.13) | -0.33<br>(-1.12, 0.46) | 0.36<br>(-0.10, 0.81) | 0.17<br>(-0.31, 0.65) | 0.71 |
| MVPA<sup>c</sup> | 0.56<br>(-0.36, 1.47) | -0.38<br>(-1.34, 0.59) | **0.67**<br>**(0.14, 1.20)** | 0.43<br>(-0.13, 1.00) | 0.84 |
| Total activity count<sup>d</sup> | 1.92<br>(-0.33, 4.16) | -0.01<br>(-2.37, 2.35) | **1.70**<br>**(0.13, 3.28)** | 1.04<br>(-0.63, 2.71) | 0.47 |
| Self-reported metrics: |  |  |  |  |  |
| Sedentary behavior<sup>c</sup> | **1.13**<br>**(0.08, 2.19)** | 0.13<br>(-0.99, 1.25) | -0.24<br>(-1.03, 0.56) | -0.48<br>(-1.29, 0.33) | 0.03 |
| All exercise-related (METs ≥2)<sup>c</sup> | **0.75**<br>**(0.06, 1.45)** | 0.21<br>(-0.50, 0.91) | **1.06**<br>**(0.52, 1.60)** | **0.86**<br>**(0.30, 1.43)** | 0.65 |
| MVPA (METs ≥3)<sup>c</sup> | **1.62**<br>**(0.62, 2.63)** | 0.85<br>(-0.18, 1.88) | **2.13**<br>**(1.27, 2.98)** | **1.76**<br>**(0.85, 2.67)** | 0.55 |

Note: Bold β were significant as P<0.05.
<sup>a</sup> The analytical samples were n=697 for maxOXPHOS. Model 1 adjusted for age and technician/site. Model 2 further adjusted for race, education, body mass index, smoking, alcohol, number of comorbidities, number of Rx medications.
<sup>b</sup> ActiGraph GT9X data were averaged over all 24 hr periods of wear (00:00 to 23:59). Sedentary behavior excluded time in-bed. Physical activity intensities were defined using Montoye et al. 2020 based on vector magnitude counts (VMC) to classify sum of total time spent (min/day) in sedentary behavior (<2860 VMC/min), light activity (2860-3940 VMC/min) and MVPA (≥3941 VMC/min).
<sup>c</sup> Re-scaled to every 30 min/day more.
<sup>d</sup> Re-scaled to 1 standard deviation = 520227 count/day more for men, 603328 count/day more for women.
Abbreviations: OXPHOS = oxidative phosphorylation; CHAMPS = The Community Healthy Activities Model Program for Seniors; MVPA =
moderate-to-vigorous physical activity; METs = metabolic equivalents.

**Supplemental Table 2.**
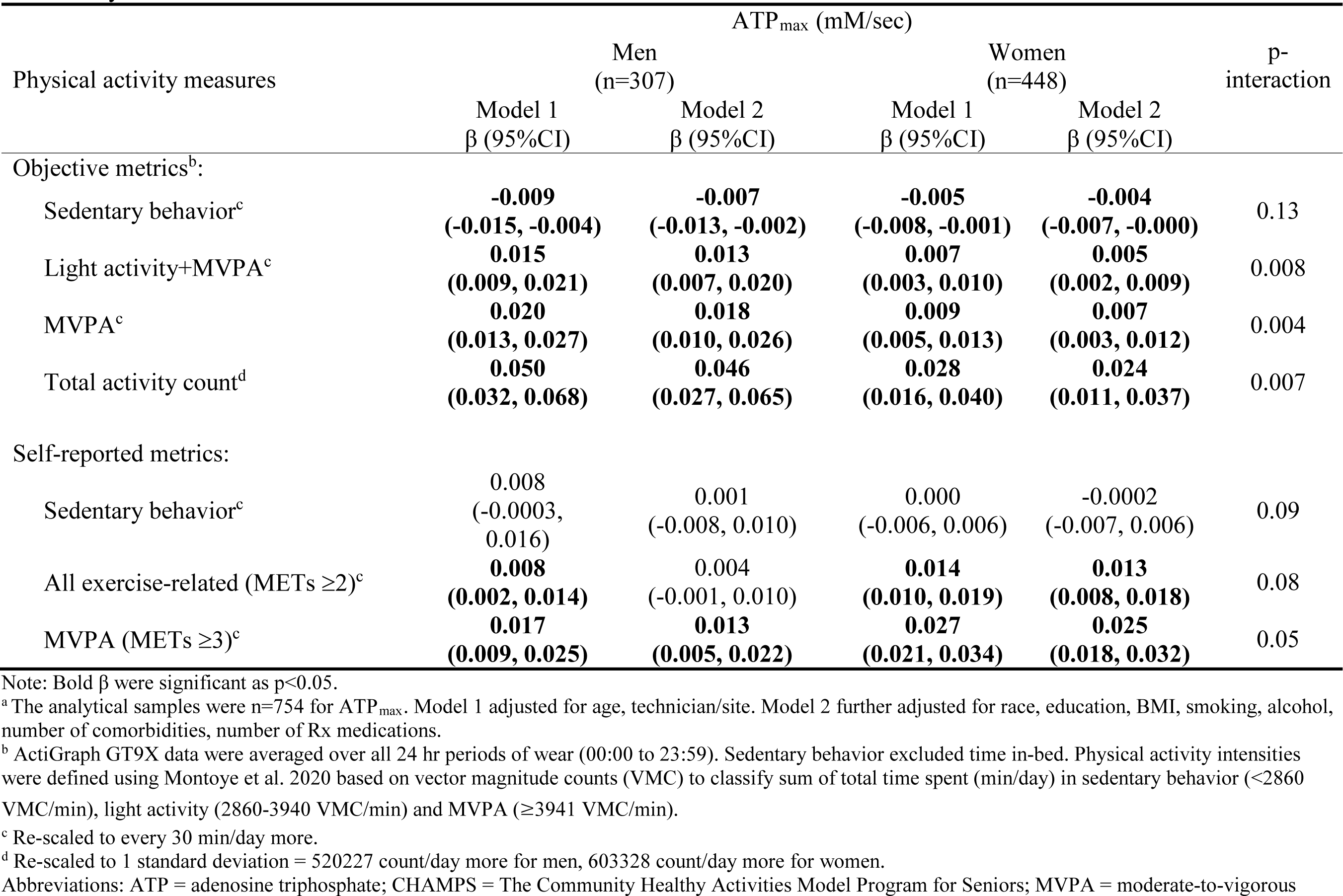
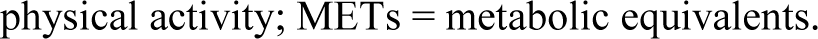
Associations of physical activity and ATP_max_ in the Study of Muscle, Mobility and Aging (SOMMA) stratified by sex^a^.

| Physical activity measures | ATP<sub>max</sub> (mM/sec) |  |  |  | p-interaction |
| --- | --- | --- | --- | --- | --- |
|  | Men<br>(n=307) |  | Women<br>(n=448) |  |  |
|  | Model 1<br>β (95%CI) | Model 2<br>β (95%CI) | Model 1<br>β (95%CI) | Model 2<br>β (95%CI) |  |
| Objective metrics<sup>b</sup>: |  |  |  |  |  |
| Sedentary behavior<sup>c</sup> | **-0.009**<br>**(-0.015, -0.004)** | **-0.007**<br>**(-0.013, -0.002)** | **-0.005**<br>**(-0.008, -0.001)** | **-0.004**<br>**(-0.007, -0.000)** | 0.13 |
| Light activity+MVPA<sup>c</sup> | **0.015**<br>**(0.009, 0.021)** | **0.013**<br>**(0.007, 0.020)** | **0.007**<br>**(0.003, 0.010)** | **0.005**<br>**(0.002, 0.009)** | 0.008 |
| MVPA<sup>c</sup> | **0.020**<br>**(0.013, 0.027)** | **0.018**<br>**(0.010, 0.026)** | **0.009**<br>**(0.005, 0.013)** | **0.007**<br>**(0.003, 0.012)** | 0.004 |
| Total activity count<sup>d</sup> | **0.050**<br>**(0.032, 0.068)** | **0.046**<br>**(0.027, 0.065)** | **0.028**<br>**(0.016, 0.040)** | **0.024**<br>**(0.011, 0.037)** | 0.007 |
| Self-reported metrics: |  |  |  |  |  |
| Sedentary behavior<sup>c</sup> | 0.008<br>(-0.0003, 0.016) | 0.001<br>(-0.008, 0.010) | 0.000<br>(-0.006, 0.006) | -0.0002<br>(-0.007, 0.006) | 0.09 |
| All exercise-related (METs ≥2)<sup>c</sup> | **0.008**<br>**(0.002, 0.014)** | 0.004<br>(-0.001, 0.010) | **0.014**<br>**(0.010, 0.019)** | **0.013**<br>**(0.008, 0.018)** | 0.08 |
| MVPA (METs ≥3)<sup>c</sup> | **0.017**<br>**(0.009, 0.025)** | **0.013**<br>**(0.005, 0.022)** | **0.027**<br>**(0.021, 0.034)** | **0.025**<br>**(0.018, 0.032)** | 0.05 |

Note: Bold β were significant as p<0.05.
<sup>a</sup> The analytical samples were n=754 for ATP<sub>max</sub>. Model 1 adjusted for age, technician/site. Model 2 further adjusted for race, education, BMI, smoking, alcohol, number of comorbidities, number of Rx medications.
<sup>b</sup> ActiGraph GT9X data were averaged over all 24 hr periods of wear (00:00 to 23:59). Sedentary behavior excluded time in-bed. Physical activity intensities were defined using Montoye et al. 2020 based on vector magnitude counts (VMC) to classify sum of total time spent (min/day) in sedentary behavior (<2860 VMC/min), light activity (2860-3940 VMC/min) and MVPA (≥3941 VMC/min).
<sup>c</sup> Re-scaled to every 30 min/day more.
<sup>d</sup> Re-scaled to 1 standard deviation = 520227 count/day more for men, 603328 count/day more for women.
physical activity; METs = metabolic equivalents.

**Supplemental Table 3.** Associations of step count and skeletal muscle energetics in the Study of Muscle, Mobility and Aging (SOMMA)^a^.

| Physical activity measures | maxOXPHOS (pmol/(s*mg)) |  | ATP<sub>max</sub> (mM/sec) |  |
| --- | --- | --- | --- | --- |
|  | Model 1<br>β (95%CI) | Model 2<br>β (95%CI) | Model 1<br>B (95%CI) | Model 2<br>β (95%CI) |
| Step counts per day (by 1000 step increase): |  |  |  |  |
| All Participants | **1.34**<br>(0.94, 1.75) | **0.97**<br>(0.54, 1.41) | **0.017**<br>(0.013, 0.020) | **0.016**<br>(0.013, 0.020) |
| Men | **1.45**<br>(0.81, 2.09) | **0.94**<br>(0.26, 1.62) | **0.018**<br>(0.012, 0.023) | **0.018**<br>(0.012, 0.023) |
| Women | **1.20**<br>(0.69, 1.71) | **1.03**<br>(0.46, 1.60) | **0.015**<br>(0.011, 0.019) | **0.014**<br>(0.010, 0.019) |
| p-interaction | **0.390** |  | **0.304** |  |

Note: Bold β were significant as P<0.05.
<sup>a</sup> The analytical samples were: All participants: n=614 for maxOXPHOS, n=660 for ATP<sub>max</sub>. Men: n=273 for maxOXPHOS, n=267 for ATP<sub>max</sub>. Women: n=341 for maxOXPHOS, n=393 for ATP<sub>max</sub>.
Model 1 adjusted for age, technician/site. Model 2 further adjusted for race, education, body mass index, smoking, alcohol, number of comorbidities, number of Rx medications. All participants models also adjusted for sex.
Abbreviations: OXPHOS = oxidative phosphorylation; ATP = adenosine triphosphate

**Supplemental Figure 1.**
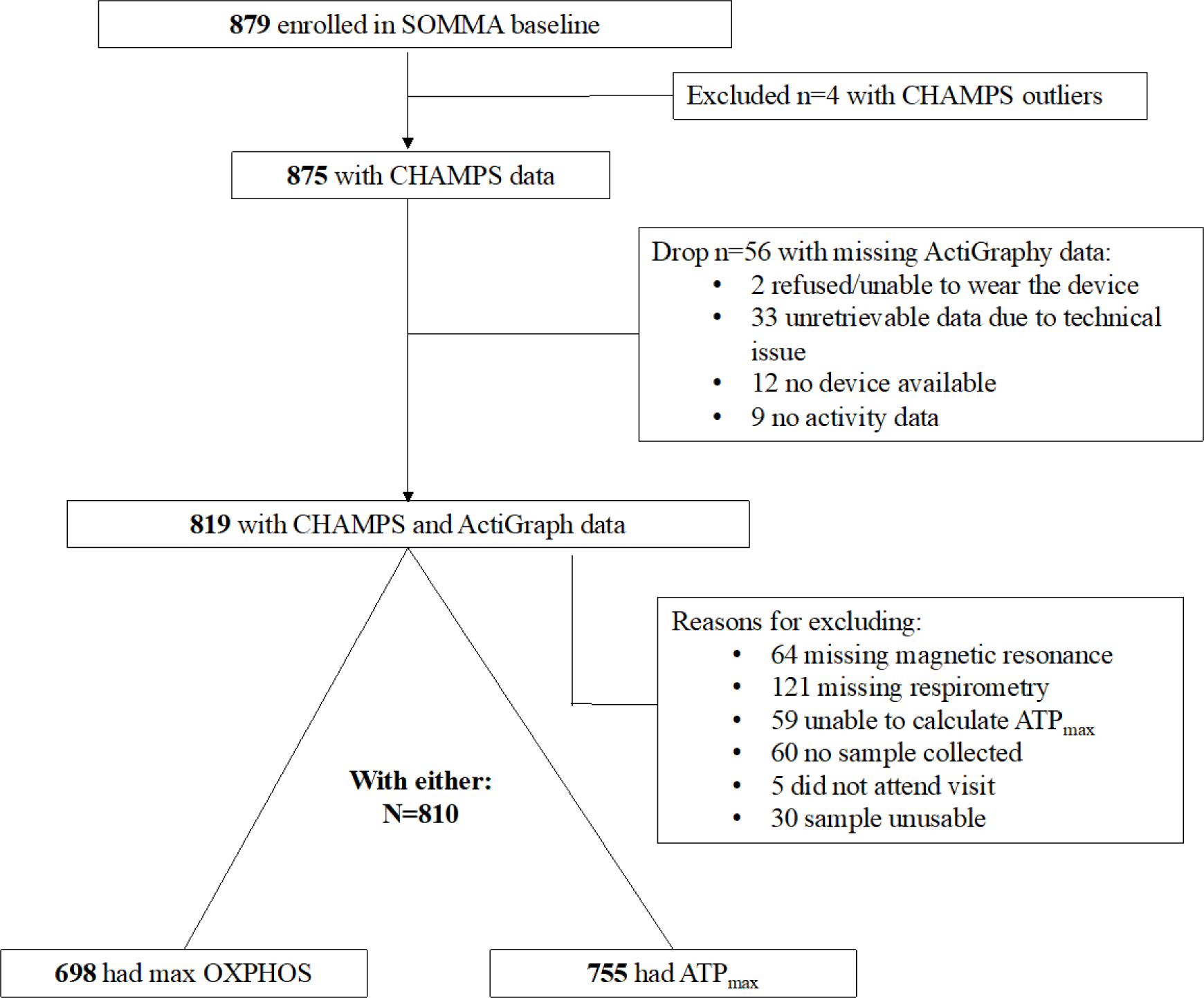
Flow chart for analytical samples in the Study of Muscle, Mobility and Aging (SOMMA) Abbreviations: ATP = adenosine triphosphate; CHAMPS = The Community Healthy Activities Model Program for Seniors; MVPA = moderate-to-vigorous physical activity; OXPHOS = oxidative phosphorylation.

